# Sex and generation shape the genetic architecture of smoking initiation

**DOI:** 10.64898/2026.11.11.26367842

**Authors:** Yeongmi Jeong, Saranya Arirangan, Evelina T. Akimova, Katherine N. Thompson, Rafael Geurgas, Jiachen Liu, Aahan Bajpai, Kathryn Fiuza Malerbi, Katelyn Kukar Bond, Julie M. Granka, Barry Hicks, David A. Hinds, Robert K. Bell, 23andMe Research Team, Geyu Zhou, Boran Gao, Mehgan Zacher, Andrew D. Grotzinger, Andrea Ganna, Michel G. Nivard, Brad Verhulst, Robbee Wedow

## Abstract

The rapidly changing cultural contexts over the past century modified the genetic factors that influence smoking initiation (SI). Using data from nearly seven million respondents, we conducted genome-wide associations study (GWAS) meta-analyses stratified by sex and generation across European, East Asian, African American, and Latino genetic ancestry populations. Sex and generation, but not genetic ancestry, moderate both the SNP-based heritability estimates and associations with individual loci. Specifically, SI is more heritable in females and younger generations. Furthermore, the impact of both discrete policy changes that target smoking and the gradual cultural redefinition of smoking norms across sex and generation are captured within the patterns of cross-trait genetic correlations between SI and extant smoking-relevant phenotypes. Robust cross-ancestry genetic correlations allowed us to meta-analyze the results across subsamples, identifying 1,717 loci associated with SI, including both main and moderated associations for either moderator. Genome-wide heterogeneity analyses identified 45 sex-specific loci and 7 generation-specific loci with distinct associations with SI. Finemapping analyses highlighted 389 and 304 loci with distinct patterns of potential causal variants by sex and generation. Accordingly, sex-and generation-specific macrosocial contexts dynamically influence the genetic architecture of SI, underscoring the need to consider gene-environment interplay for health behaviors as societal norms evolve.

## Introduction

Tobacco use remains one of the leading preventable causes of premature mortality worldwide, accounting for an estimated 8.7 million deaths annually.^1,2^ Since the 1964 Surgeon General’s Report on Smoking and Health^1^, public policies and shifting cultural norms have contributed to a substantial decline in smoking prevalence rates in the general population. Nevertheless, smoking initiation (SI) has not declined uniformly across sub-populations: SI is higher in males and older generations.^3^ These subpopulation differences may mirror differences in the genetic architecture of SI in particular groups, resulting in partially distinct genetic associations, variable heritability estimates, and differential patterns of genetic correlation across a range of outcomes. Within a gene-environment interaction (GxE) framework, a person’s sex or the historical context of their lives can enable, constrain, or redirect their genetic propensity for smoking. However, little work has integrated these macro-level transformations into well-powered, molecular genetic models of SI. For example, the Surgeon General’s report may have shifted the likelihood of SI for individuals with some genotypes, changing the pattern of genetic associations for those who initiated before or after the report. Similarly, gradual changes in the stigmatization of smoking behaviors, as well as gendered norms for substance use, can induce associations between SI and clusters of sociodemographic, psychological, and medical phenotypes.^4^

Converging evidence across twin and family studies^2,5–7^, genome-wide association studies^4,8^, polygenic risk scores^4,8^, and rare variant burden analyses emphasize the integral role of genetic factors in SI. However, the genetic components of SI operate within an individual’s environmental context. The heritability of smoking varies across sex^9^, birth cohort^10,11^, and location^12^, suggesting genetic influences may become more or less salient depending on the normative and regulatory environment surrounding tobacco use when people enter the developmental risk window for SI.^10^ Such variation in the magnitude of the genetic effects could arise when contextual factors make it more difficult or costly to engage in SI, raising the threshold required to initiate. As a consequence, SI could become increasingly concentrated among individuals with heritable traits such as impulsivity (a key dimension of ADHD) or risk tolerance. This implies that effect sizes derived from GWAS reflect specific regulatory and normative contexts at the time of initiation.

We focus on two environmental contexts: sex and generation. We begin by examining differences in single nucleotide polymorphism (SNP)-based heritability across sex and generations. We also perform locus-level analyses on sex and generation, using heterogeneity in the effect sizes and subgroup specific patterns of candidate causal variants as evidence of GxE. Our work extends previous twin and family research by using genome-wide data from nearly 7 million individuals in an ancestrally diverse sample collected by the 23andMe Research Institute, a nonprofit medical research organization. The historic magnitude of this ancestrally diverse data allows us to demonstrate the contextual importance of sex and generation in shaping the genetics of SI. By embedding genomic analyses within a life course and sex differences framework, we offer a more complete account of smoking initiation as both a biological and social phenomenon, one that illustrates how integrative models of GxE can deepen understanding of health behaviors as societal norms evolve. SI is a particularly compelling phenotype to explore GxE, given the major environmental changes that have occurred over the last century.

## Results

### Genomic Mechanisms of Sex- and Generational Moderation of SI

SI was defined as a binary outcome indicating whether the participant had smoked at least 100 cigarettes in their lifetime. Stratified GWAS analyses of SI were conducted for a) sex (N_males_ = 2,575,611, N_females_ = 3,206,596, N_Total Sex_ = 5,782,207), and b) 16 birth cohorts roughly reflecting four generations (N_Silent Generation (1924-1943)_ = 405,363, N_Baby Boomers (1944-1963)_ = 1,906,551, N_Generation X (1964-1983)_ = 2,314,494, and N_Millennials (1984-2003)_ = 2,325,158, N_Total Generation_ = 6,951,566) using data provided by 23andMe Research Institute (see **Supplementary Table S1** for details). For simplicity, we used colloquial naming conventions for the generations despite minimal deviations between the birth years (see **Box 1** in **Methods**). For each moderator, stratified analyses were conducted within four continental ancestry groups: European-(EUR), African American-(AFR), Latino-(AMR), and East Asian-like (EAS) ancestries. Samples are screened for relatedness to ensure only one family member is included per subsample, but allowing multiple family members to contribute to different sex or birth cohorts. Thus, N_Total Sex_ is smaller than our N_Total Generation_ number, since more family members could be split into the 16 independent birth cohort-stratified GWAS than could be split into the 2 sex-stratified GWAS.

#### SNP-Based Heritability (h^2^_snp_)

We used LD-Score Regression (LDSC)^13^ to estimate h^2^_snp_ and cross-trait genetic correlations (rG_snp_) in each sex and generation. As LDSC was developed using EUR-like ancestry LD patterns, and most existing smoking-relevant GWAS analyses have been conducted in EUR-like samples, we focus on the EUR-like ancestry summary statistics (**Table S1**). As a starting point, in the non-stratified sample, h^2^_snp_ of SI is 0.0849 (standard error [se]=0.0019: **Figure 1**). This pooled estimate, however, masks the fact that SI is significantly more heritable among females (h^2^_snp_=0.0951; se=0.0022) relative to males (h^2^_snp_=0.0854; se=0.0021; *p_(female=male)_*=0.0014; see **Figure 1a**). Further, the h^2^_snp_ of SI increases steadily across generations, with h^2^_snp_ of SI in the Silent Generation of 0.0813 (se=0.0033), Baby Boomers of 0.0868 (se=0.0022), Generation X of 0.0924 (se=0.0023), and Millennials of 0.0944 (se=0.0023; see **Figure 1b**). Estimates across the 16 five-year birth cohort bins show a similar pattern. After multiple testing corrections (adjusted *p*=0.0083) pairwise Wald tests indicate that h^2^_snp_ for the Silent Generation is significantly lower than Generation X (*p*=0.0058) and Millennials (*p*=0.0011). The gradual nature of the generational change is emphasized by the fact that differences between adjacent generations do not survive correction for multiple testing, nor do any differences with the Baby Boomers (see **Supplementary Table S3** for all test results). These results suggest genetic factors play a larger role in SI for females and younger generations. (See **Supplementary Table S2** for the h^2^_snp_ of SI across the sixteen five-year birth cohorts.)

We also estimated h^2^_snp_ for the five US regions that have historically shown differences in smoking policies and behaviors (N_midwest_ = 844,743, N_northeast_ = 833,087, N_southeast_ = 1,022,535, N_southwest_ = 566,432, and N_west_ = 1,247,231, N_Total Regions_ = 4,514,028; **Tables S4** and **S5**). These estimates were limited to individuals living in the US. We observed no evidence of meaningful regional heterogeneity in the heritability of SI and therefore drop region from our locus-level analyses.

#### Genetic Architecture of SI across Levels of Sex and Generation

We estimated rG_snp_ between a) females and males (**Figure 1a**) and b) the different generations (**Figure 1b**) to assess the overlapping genetic architecture of SI across the levels of sex and generation. The genetic correlation for SI between males and females is 0.887 (se=0.0046; *p_(rG=1)_=*2.97e-133), indicating that while the majority of the genetic influences are shared across sexes, SI has a partially sex-specific genetic architecture. Similarly, we observe a clear gradient in the genetic correlations across generations, with larger genetic correlations between adjacent birth cohorts (rG_snp_=0.94-0.99) that decline steadily as temporal distance increases (rG_snp_=0.83-0.94). Genetic correlations across all 16 birth cohorts are presented in **Figure 1b**, with a similar pattern of genetic correlation across generations (rG_snp_=0.62-0.98). Thus, slightly different genetic factors play a role in the genetic architecture of SI based on both the person’s sex or generation. (For the near unity correlations for the region-stratified analyses, see **Figure S1**).

#### Cross-Trait Genetic Correlations with External Phenotypes

To establish a baseline for comparing the moderated genetic correlations, we estimated cross-trait rG_snp_ between the unmoderated meta-analysis and a curated set of relevant phenotypes (**Figure 1c**). The baseline pattern of correlations is consistent with previous genetic correlations between SI and other substance use, psychiatric, and personality traits, socioeconomic and health-related outcomes.^4^

Across generations, we observed two distinct patterns of cross-trait genetic correlations. The first pattern revealed the Silent Generation had the most divergent pattern of associations relative to the subsequent three generations. This is consistent with an acute historical time effect: the 1964 Surgeon General’s report^1,14^ was issued when the Baby Boomers were at the approximate age to start smoking. Thus the magnitude of the genetic correlations between SI and age of initiation (rG_Silent Generation_ = -0.57; rG_Others_ ≈ -0.68), cigarettes per day (rG_Silent Generation_ = 0.15; rG_Others_ ≈ 0.30), and smoking cessation (rG_Silent Generation_ = 0.24; rG_Others_ ≈ 0.35) are significantly smaller in the Silent Generation relative to subsequent generations. Similarly, the magnitude of genetic correlations with educational attainment increased markedly (rG_Silent Generation_ = -0.25; rG_Others_ ≈ -0.40). As such, one possible interpretation of our results is that the Surgeon General’s report triggered a relatively abrupt shift in nicotine use behaviors, particularly for more educated populations.

The second pattern of genetic correlations is consistent with a gradual cultural redefinition of smoking norms. Specifically, the magnitude of the cross-trait rG_snp_ increased across several sociodemographic factors, and psychiatric and medical phenotypes in the younger generations. From a social determinants of health perspective, SI was correlated with lower rates of satisfaction with familial and financial aspects of a person’s life, an increased likelihood of living in disadvantaged neighborhoods, and less frequent religious attendance. For the psychiatric and medical phenotypes, SI was increasingly correlated with general risk tolerance, loneliness, and neuroticism, and the psychiatric disorders schizophrenia, ADHD, MDD and bipolar disorder in younger generations. Thus, over the course of 4 generations, the magnitude of genetic correlations between SI and psychiatric traits and disorders slowly and significantly changed.

We then applied autoregressive latent change score models in Genomic Structural Equation Modeling^15^ that estimated the genetic overlap between external traits and the generation-specific component of the genetic signal in SI that remains when removing shared signal with prior generations (**Supplementary Note B**; **Figures S3** and **S4**). These results recapitulate the joint shift in the SI risk factors and the sociocultural forces that shape smoking over time. Specifically, a wide array of genetic correlations with generation-specific signals in SI were significantly different from zero, highlighting the value of generation stratified analyses that would otherwise be distorted or diluted when collapsing across generations. We highlight two findings here. When removing shared genetic signals with the Silent Generation, the generation-specific signal for Baby Boomers was positively correlated with ADHD at 0.22. Similarly, the generation-specific genetic signal for SI in Millennials was positively correlated with genetic signal for risk tolerance. These results could indicate that as societal attitudes and health norms have increasingly discouraged smoking behaviors, that a corresponding increase in genetic liability for disinhibited traits is required to engage in smoking over time.

We also observed differential genetic correlations with several external phenotypes based on sex (**Figure 1c**). Note that tests of differences between sexes are indicated in **Figure 1c** in the *δsex* column. Specifically, SI is more strongly genetically correlated with smoking cessation in females (rG_Females_ = 0.39, rG_Males_ = 0.31). For the sociodemographic traits, SI in females was more strongly correlated with living in more disadvantaged neighborhoods (rG_Females_ = 0.62, rG_Males_ = 0.54), and lower rates of church attendance (rG_Females_ = -0.42, rG_Males_ = -0.35), emphasizing how genetic factors may underscore the relationship between SI and standard social determinants of health. When examining the psychiatric and medical phenotypes, SI was more genetically correlated with higher BMI (rG_Females_ = 0.24, rG_Males_ = 0.19), and increased likelihood of ADHD (rG_Females_ = 0.66, rG_Males_ = 0.58), and higher rates of risk tolerance (rG_Females_ = 0.34, rG_Males_ = 0.27) in females. Notably, the moderated genetic correlations in sex and across generational groups were not entirely consistent, implying each moderator regulated slightly different pathways that culminate in SI.

**Figure 1.**
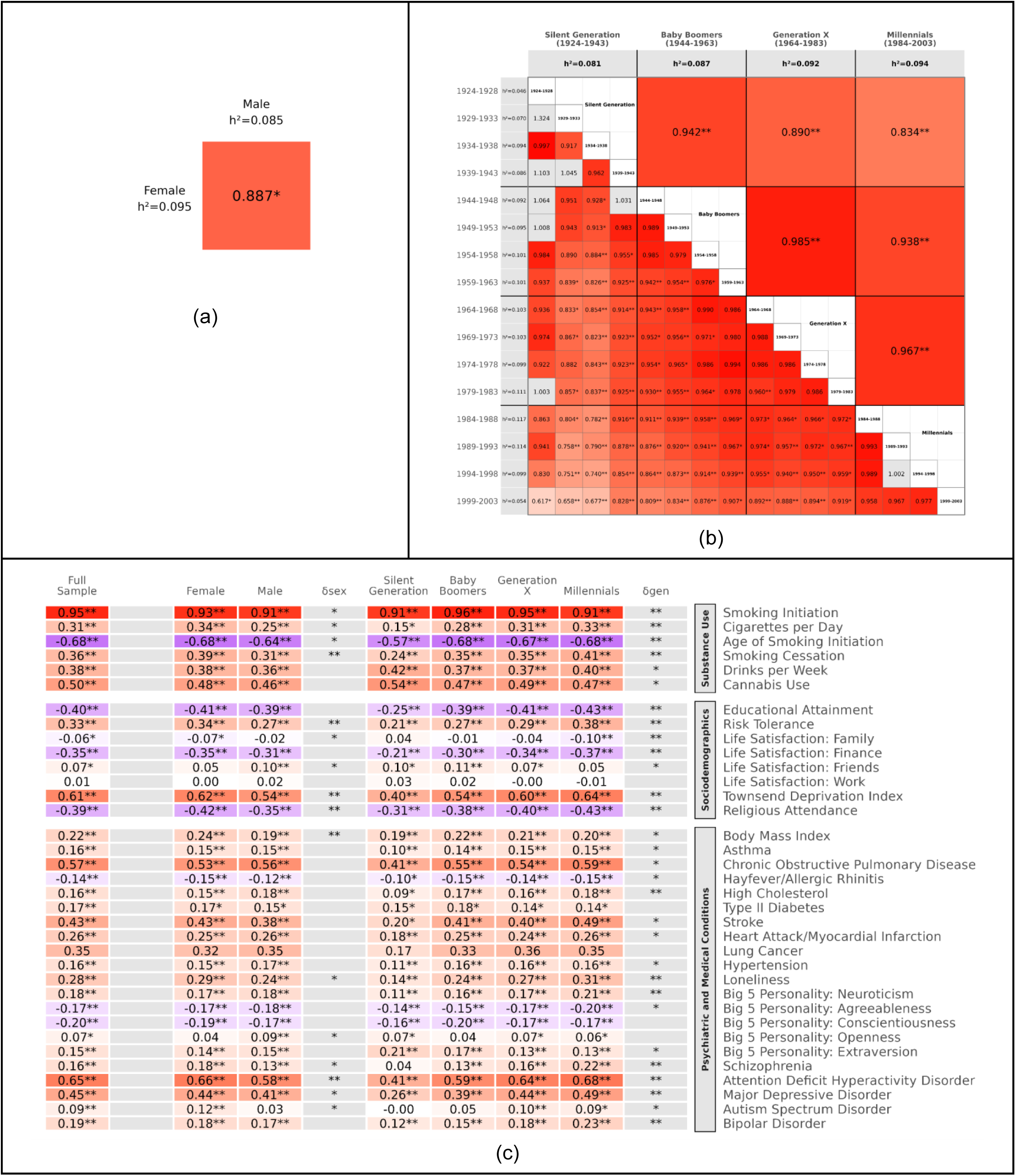
SNP Heritability of Smoking Initiation, Genetic Correlations across Sex and Generation Subgroups, and Cross-Trait Genetic Correlations with substance use, sociodemographic, and psychiatric and medical conditions. Panels a and b show SNP heritability (h^2^_snp_) and within-trait genetic correlations (rG_snp_) of smoking initiation across sex and generations. Panel c presents cross-trait genetic correlations between smoking initiation and other substance use, sociodemographic, and psychiatric and medical conditions. Cross-trait genetic correlations for all 16 birth cohorts are presented in Supplementary Figure S2. All h^2^_snp_ and rG_snp_ estimates were obtained using LD score regression in EUR-like ancestry summary statistics. Purple shading indicates genetic correlations close to negative one, whereas red shading denotes genetic correlations close to one. Darker shading reflects stronger correlations. In panels a and b, a single asterisk (*) denotes a genetic correlation significantly different from 1 at *p < 0.05*. Double asterisks (**) indicate significance after Bonferroni correction: *p < 0.000416* (corrected for 120 independent tests) for the birth cohort analyses and at *p < 0.0083* (corrected for 6 independent tests) for the generation analyses. For sex comparisons, no multiple-testing correction was applied because only one hypothesis test was conducted. In panel c, a single asterisk (*) indicates a genetic correlation significantly different from 0 at *p < 0.05*. Double asterisks (**) indicate significance after Bonferroni correction: *p < 0.0014* for the full sample (corrected for 35 independent tests), *p < 0.0007* for the sex-stratified sample (corrected for 70 independent tests), and *p < 0.0004* for the generation sample (corrected for 140 independent tests). We report pairwise equality tests of genetic correlations within each moderator (i.e., sex and generation) in the columns labeled *δsex* and *δgen*. In the *δsex* column, a single asterisk (*) indicates that the genetic correlation differs significantly between females and males at *p < 0.05*, whereas double asterisks (**) indicate significance after Bonferroni correction at *p < 0.0014* (corrected for 35 independent tests). In the *δgen* column, a single asterisk (*) indicates that the genetic correlation differs significantly for at least one pair of generations at *p < 0.05*, and double asterisks (**) indicate significance after Bonferroni correction at *p < 0.0002* (corrected for 210 independent tests). Complete pairwise test results are provided in Supplementary Table S6. SI in row 1 of panel c comes from a previous meta-analysis of this phenotype using different samples.^4^

### Multi-Ancestry Locus-Level Heterogeneity Analyses

We conducted a series of meta-analyses across sex and birth cohort using inverse variance weighting with METAL within each ancestry.^16^ The Popcorn software package^17^ estimated moderate to high cross-ancestry genetic effect correlations at each level of the moderators (**Supplementary Note C**), suggesting ancestry did not modify the genetic architecture of SI (**Figure S5**). Accordingly, we present pan-ancestry meta-analyses for the locus-level results. To be considered significant moderation, a variant had to meet two criteria: (i) the SNP had to demonstrate genome-wide significant heterogeneity across generations or sexes (*HetPVal* < 5 × 10^−8^) and (ii) it had to be genome-wide significant (*p* < 5 × 10^−8^) in at least one subgroup. Significant main effect variants had to have a genome-wide significant association in at least one subgroup, without genome-wide significant heterogeneity. **Figure 2** presents a Manhattan plot of the full meta-analysis results, highlighting lead SNPs with significant heterogeneity (i.e., moderation) in the sex and birth-cohort meta-analyses in red and green, respectively. In total, we identified 1,717 meta loci for SI across both the sex and generation analyses, including 45 loci that were moderated by sex and 7 loci that were moderated by generation. These analyses suggest that while the genetic architecture of SI was broadly stable across a person’s sex and generation, several notable exceptions underscore the contextual sensitivity of specific genetic loci.

**Figure 2.**
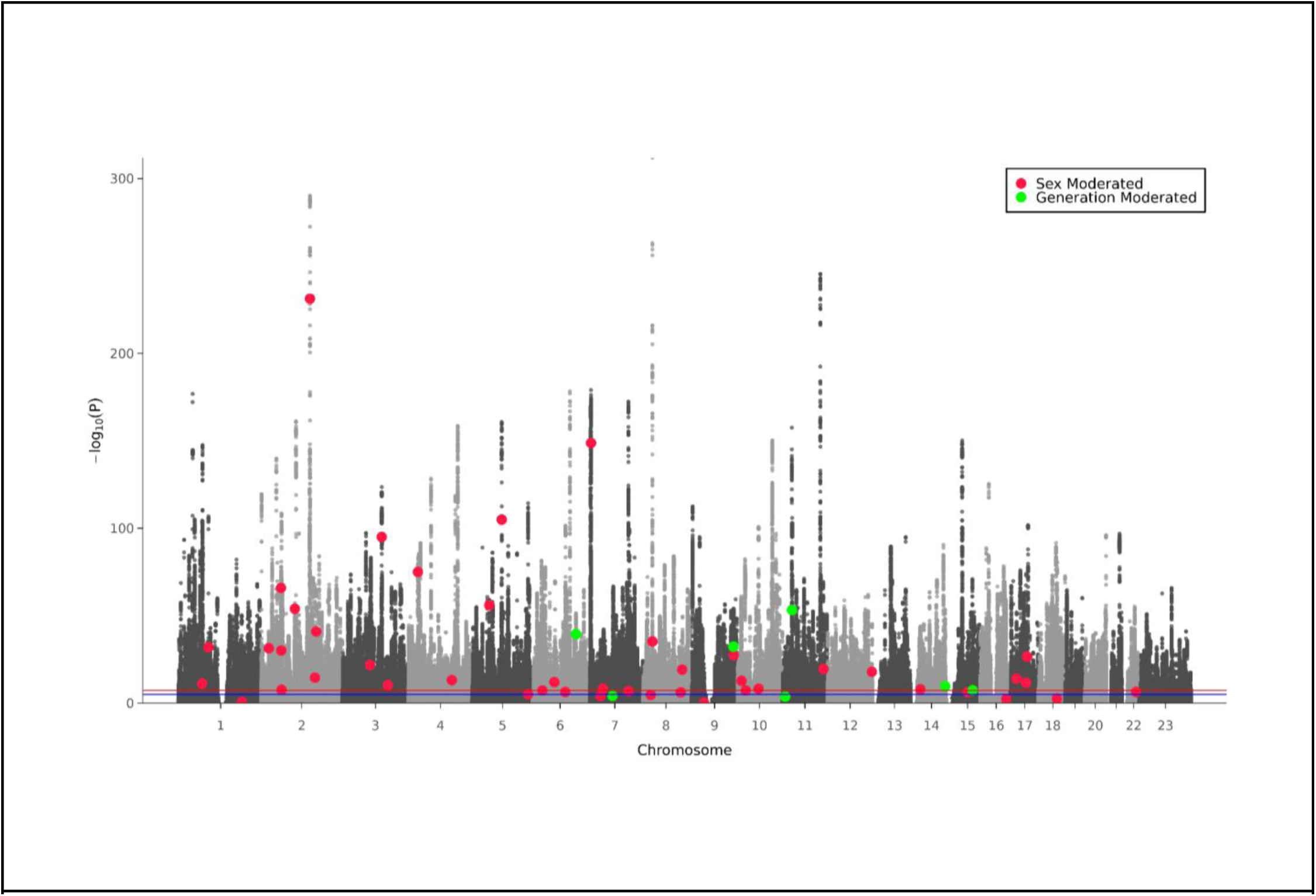
Manhattan Plot of Meta-Analytic Heterogeneity. The x-axis shows the position in the genome (chromosomes 1 to 22 and X) and the y-axis represents −log10(P values) (two-sided) for the association of variants with SI using an inverse-variance weighted fixed effects model (2,237,272 cases and 2,664,582 controls). The horizontal red line shows the threshold for GWAS (P = 5 × 10−8). Dots represent each SNP that was tested in the GWAS. To highlight genomic moderation, red and green dots indicate the lead SNPs of loci with significant heterogeneity across levels of sex and generation, respectively.

#### Variant-Level Moderation by Generation

For the 7 meta loci moderated by a person’s generation, the heterogeneity was most pronounced in comparisons involving the earliest (Silent Generation) and the most recent (Millennials) generations. **Figure 3** presents two selected SNP-by-generation effects that demonstrate distinct patterns of locus level genomic moderation for SI. **Figure 3a** illustrates moderation of the well-established CHRNA5/3 locus on chromosome 15. The strong, significant association with SI in the Silent Generation is substantially attenuated in both Millennials (**Figure 3a**) and Generation X (**Figure S6a**). In contrast, the SCAI loci on chromosome 9 demonstrate emergent or amplified effects in Millennials relative to the Silent Generation (**Figure 3b**). The contrasting patterns indicate that genetic associations may become more salient under contemporary smoking environments. These findings are consistent with cohort-specific moderation of genetic pathways potentially related to behavioral regulation or biological responses to nicotine exposure. Analogous figures for the remaining moderated meta loci identified in the SNP-by-generation analyses are presented in **Supplementary Figure S6**.

**Figure 3.**
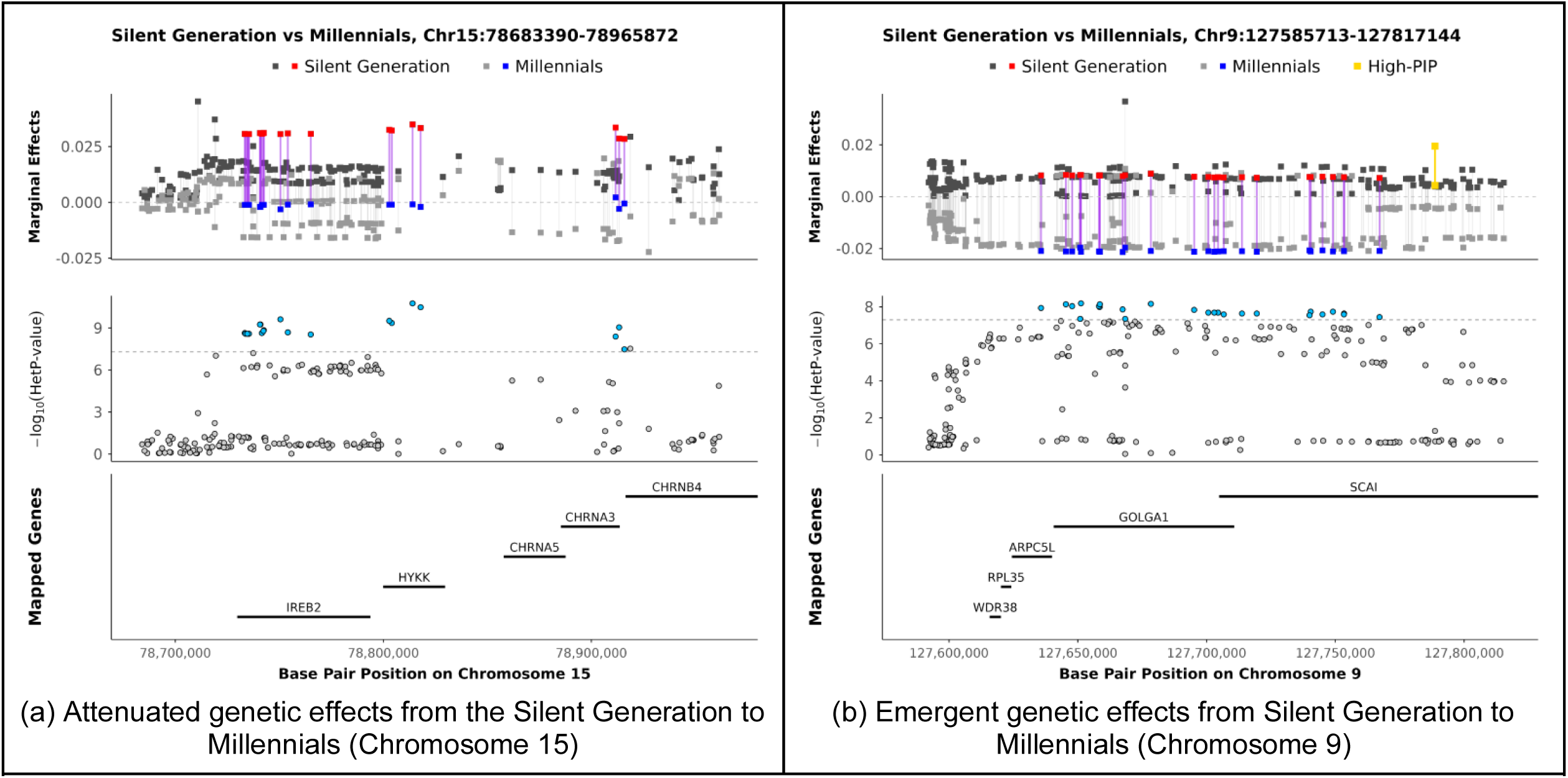
Locus-Specific Heterogeneity in Genetic Effects across Generations on Chromosome 15 and 9. Panels a and b present locus-specific heterogeneity in SNP effects between the Silent Generation and Millennials for two moderated effect meta loci. Panel a shows the CHRNA5/3 locus on chromosome 15 , where the genetic effects observed in the Silent Generation are attenuated among Millennials. Panel b shows the SCAI locus on chromosome 9 exhibiting an emergent moderation pattern, with genetic effects becoming more pronounced among Millennials than in the Silent Generation. In each panel, the top plot displays the difference in SNP effect size estimates for the two generations. The red/blue dots indicate variants with GWAS significant heterogeneity (Het P-value) between the Silent Generation (red) and the Millennials (blue) while the grey-scale dots indicate effect sizes for non-significant het P-values. The middle plot displays the corresponding heterogeneity p-values. The bottom plot shows positionally mapped genes within the locus. The panel title reports the chromosome and the start and end base-pair positions of the displayed locus. SNP positions are based on the GRCh37/hg19 genome build. High-PIP variants from the fine-mapping analyses are shown in yellow only when they fall within the boundaries of the displayed locus; variants outside these boundaries are not shown.

As a complementary approach to assess temporal heterogeneity in genetic effects, we examined linear trends in SNP effect sizes across the 16 5-year birth cohort bins using generalized least squares (GLS) regression (**Supplementary Note E**). None of the 1,670 unique SNPs within the main effect meta loci exhibited significant temporal trends after Bonferroni correction for 1,670 tests (*p* < 0.0000299) (**Figure S8**; **Table S7**). In contrast, six of the seven unique SNPs within the seven moderated effect meta loci exhibited significant linear trends after Bonferroni correction for seven tests (*p* < 0.00714) (**Figures S9** and **S10**; **Table S8**). These findings provide complementary evidence that heterogeneity in the moderated loci is accompanied by systematic temporal changes in genetic effects across generations (**Supplementary Note E)**.

#### Variant-Level Moderation by Sex

We identified 45 meta loci with statistically significant heterogeneity across sexes distributed across multiple chromosomes. Some loci exhibit stronger associations in males (e.g., **Figure 4a**) and others in females (e.g., **Figure 4b**). Analogous figures for the remaining moderated meta loci identified in the SNP-by-sex analyses are presented in **Figure S7**. The observed sex differences may reflect a combination of biological mechanisms, such as sex-specific gene expression, as well as socially mediated differences in smoking-related environments and behaviors.

**Figure 4.**
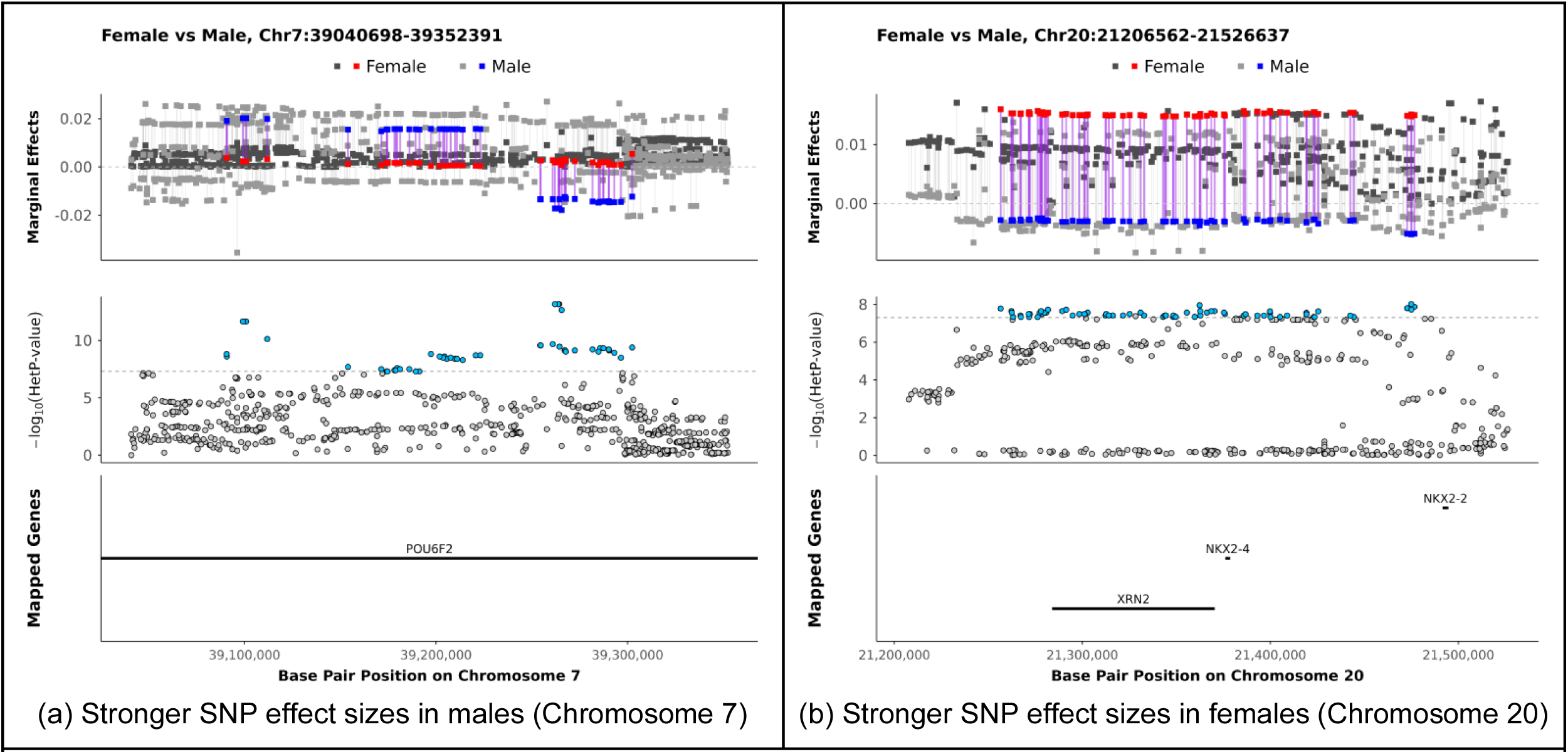
Locus-Specific Heterogeneity in Genetic Effects across Sex on Chromosomes 7 and 20. Panels a and b present locus specific heterogeneity in SNP effects between females and males for two moderated effect meta loci. Panel a shows the POU6F2 locus on chromosome 7 exhibiting stronger genetic effects among males than females. Panel b shows the XRN2 locus on chromosome 20 exhibiting stronger genetic effects among females than males. In each panel, the top plot displays SNP effect size estimates for females and males. The red/blue dots indicate variants with GWAS significant heterogeneity (Het P-value) between females (red) and males (blue) while the grey-scale dots indicate effect sizes for non-significant het P-values. The middle plot displays the corresponding heterogeneity p-values. The bottom plot shows positionally mapped genes within the locus. The panel title reports the chromosome and the start and end base pair positions of the displayed locus. SNP positions are based on the GRCh37/hg19 genome build. High-PIP variants from the fine-mapping analyses are shown only when they fall within the boundaries of the displayed locus; variants outside these boundaries are not shown.

#### Fine Mapping Analysis

We performed statistical fine-mapping using the Sum of Single Effects (SuSiE) method^18,19^ within the 1,717 genome-wide significant main effect and moderated association regions identified from the sex and generation stratified analyses. High-confidence fine mapped variants were defined as SuSiE PIP ≥ 0.8. Distinct high-PIP variants for one generation or sex implies that potential causal variants are context specific. By contrast, high-PIP variants that are consistent across contexts implies a robust potential causal signal. For sex, 268 loci only had high PIP values in females (262 main effects and 6 moderated effects), 121 only in males (116 main effect and 5 moderated effects), and 294 loci (267 main effect and 27 moderated effects) had high PIP values that were shared across sexes. For the generation finemapping analyses, 2 loci had high PIP values only in the Silent Generation, 80 had high PIP values loci only in Baby Boomers, 128 loci had high PIP values only in Generation X, and 94 loci had high PIP values only in Millennials. Thus, 389 loci had sex-specific patterns of high-PIP variants and 304 had generation-specific patterns **(Supplementary Table S9)**. Notably, the median absolute difference in PIP values between sexes and generations was relatively large (**Δ** PIP > .47), emphasizing the magnitude of the difference between potential causal variants for a specific sex or generation. For the moderated loci, comparisons between the PIP values for sex are presented in **Supplementary Table S10** and for generation are presented in **Supplementary Table S11**.

Interestingly, 683 of 1,529 sex main effect meta loci contained at least one high-PIP variant, while 38 of 45 sex-moderated meta loci contained at least one high-PIP variant **(Supplementary Figure S11)**. Similarly, 599 of 1,670 generation main effect meta loci and 5 of 7 generation moderated meta loci contained high PIP variants **(Supplementary Figure S12)**. As such, proportionally moderated loci were more frequently supported by localized fine mapping evidence than main effect loci.

#### Integrating GWAS Heterogeneity and Finemapping Analyses across Moderators

To delve into the nuanced patterns of genomic moderation, for each of the 1,717 meta-locus, we examined the overlap between the sex-and generation heterogeneity loci for main and moderated effects from the GWAS meta-analyses and the shared and specific patterns of potentially causal variants. Overwhelmingly, the main effects overlap across generation and sex, but the moderated effects in the sex analyses tended to be main effects in the generation analyses, and vice versa (**Table S12, Figure 5a**). Only one locus was moderated by both sex and generation. Similarly, for the finemapping analyses, of 541 high-confidence finemapped regions, the plurality of loci (210) had the same pattern of potentially causal variants across both sex and generation. However, for the finemapped analyses, several loci (18) shared both sex-and generation-specific patterns of potentially causal loci (**Table S12, Figure 5a**).

Looking at the intersection of main and moderated GWAS effects with the pattern of finemapped variants within either sex (**Figure 5c**) or generation (**Figure 5d**), main effect loci tended to share the same pattern of potentially causal variants across levels of the moderators. However, several main effect loci had either sex-(116 loci) or generation-specific (94 loci) patterns of potentially causal variants. When examining the moderated loci with high confidence finemapped variants, the same pattern of potentially causal variants tended to emerge, despite differences in the magnitude of the GWAS effect sizes.

Finally, we evaluated the meta loci for both GWAS heterogeneity and differential patterns of finemapped variants across both moderators (**Figure 5e**). Most notably, 180 loci had main effects for both moderators and a shared pattern of high confidence variants in both the sex and generation. Another 131 loci had both sex and generation main effects, but a high confidence variant for one of the moderators. This pattern of results implies strong and unambiguous evidence of genomic main effects. The remaining 597 loci displayed some evidence of contextual sensitivity by showing significant sex or generation heterogeneity in the GWAS meta-analyses or distinct patterns of potential causal loci at a specific level of the moderators (red bars).

**Figure 5.**
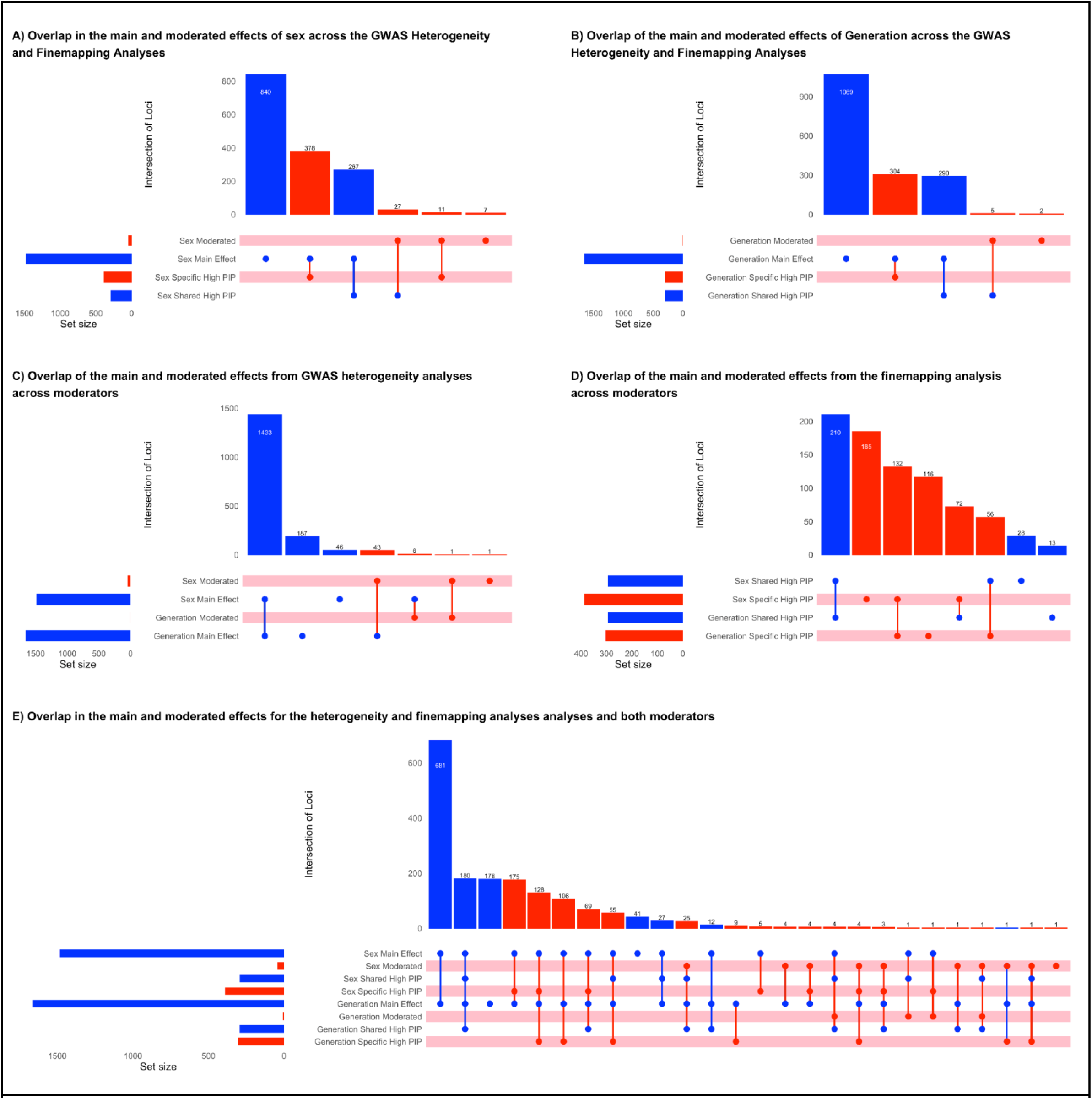
Integration of GWAS Heterogeneity and Fine-Mapping Results across Sex and Generation. Summary of the overlap of meta loci identified from GWAS heterogeneity analyses and fine-mapping analyses across sex and generation. Panels a and b compare main and moderated effect meta loci across sex and generation, respectively. Panels c and d integrate GWAS main and moderated effect meta loci with patterns of high-confidence fine-mapped variants within sex and generation, respectively. Panel e summarizes GWAS main and moderated effect meta loci as well as the fine-mapping patterns across both sex and generation. In each UpSet plot, horizontal bars indicate the total number of meta loci in each set, vertical bars indicate the number of meta loci corresponding to each intersection, and connected dots below the vertical bars identify the sets included in each intersection. Shared high-PIP indicates meta loci with a shared pattern of high-confidence fine-mapped variants across levels of the moderator, whereas specific high-PIP indicates meta loci with moderator-specific patterns of high-confidence fine mapped variants. High-confidence fine-mapped variants are defined as variants with PIP ≥ 0.8. Across all panels, the blue bars indicate main effects and the red bars indicate moderated effects. The pink bands within each figure indicate potential moderated effects. Loci with evidence for both main and moderated effects are treated as moderated effects.

## Discussion

Using multi-ancestry genome-wide data from nearly seven million individuals, we demonstrate that the genetic architecture of smoking initiation (SI) varies across sex and generation. At the locus level, we identified 1,717 loci associated with SI across both moderators, of which 1,120 were main effects and 597 were moderated effects. The moderated effects could be broken down into 45 loci exhibiting heterogeneity by sex and 7 with heterogeneity across generations. Further, 389 loci had distinct patterns of potential causal variants by sex and 304 loci had distinct patterns by generation (**Figure 5**). At the genome-wide level, we observed higher SNP-based heritability among females and younger generations. Consistent with the locus level moderation results, there were strong but imperfect genetic correlations across the sexes and generations, suggesting broadly overlapping associations with a subset of variants that differentially contribute to SI in females relative to males, and older versus younger generations. Furthermore, cross-trait genetic correlations emphasize how the changing gender and generational normative contexts alter who starts smoking (**Figure 1**). These results, utilizing distinct methodological approaches, advance our understanding of how contextual factors shape the genetic architecture of SI.

When looking across analyses, the main effects tend to have consistent impacts on SI across generations, sexes, regions, and ancestries. These associations provide the foundation for the genetic architecture of SI across populations and are the focus of most genomic research in SI.^4,8^ The moderated effects, by contrast, provide a more nuanced, contextualized perspective on genomic risk. For example, in the Silent Generation we observe a strong, significant association near the cluster of nicotine receptor genes on chromosome 15 (i.e., *CHRNA5*, *CHRNA3*, *CHRNB4*) that is modest and non-significant in Millennials (**Figure 3a**) and Generation X (**Supplemental Figure S6**). Similar patterns of amplification/suppression are observed for numerous genomic loci for both sex and generation (**Supplemental Figures S6 and S7**). Thus, sex differences and historical changes reshape associations with SI for multiple genetic variants within each locus. This implies that public health initiatives and cultural changes not only shifted SI at the population level, but these macro factors shifted associations within localized genomic regions.

A complementary and novel perspective for the genomic moderation of SI is provided by finemapping analyses.^18,19^ We identified 389 sex-specific high-confidence candidate variants and 304 generation-specific high-confidence candidate variants. These analyses leverage the distinct patterns of LD across ancestries, further emphasizing the importance of including diverse populations in GWAS analyses. Observing distinct patterns of high-confidence candidate variants in females versus males, or for different generations, implies differences in the underlying genomic architecture which is the hallmark of GxE. Importantly, genetic associations that appear stable across sex and generation can nevertheless implicate different potentially causal variants. Notably, moderated loci were proportionally more likely than main effect loci to yield high-confidence fine mapping signals, further highlighting the complementary value of these approaches. The finemapping results emphasize that locus-level moderation of SI may extend beyond differences in effect sizes to differences in the putative causal variants themselves. Integrating fine mapping and GWAS heterogeneity analyses reveals a more nuanced picture of genomic moderation than either approach provides alone. Together, these findings demonstrate that the lack of heterogeneity in the genetic associations does not necessarily imply stability in their underlying architecture, just as differences in association strength need not reflect differences in the implicated causal variants.

The genome-wide heritability and genetic correlation analyses provide an alternative perspective on GxE for SI. As LDSC does not require individual variants to be genome-wide significant, h^2^_snp_ analyses are able to incorporate the subtle signals that are missed at the level of individual genetic variants, highlighting substantially more moderation in the genetic architecture of SI than genome-wide significant loci. We observed higher heritability estimates among females and younger generations, indicating common genetic variants play a larger role in smoking initiation in these groups.

The genetic correlations extend the heritability results, illustrating how both historical events and gradually changing gendered expectations and changing social norms restrict the types of people who initiate. Consistent with a life-course and historical perspective, our generation analyses suggest that population-level policy changes, such as the 1964 Surgeon General’s report, altered patterns of smoking behaviors. Specifically, we observed particularly stark differences in the genetic correlations between the Silent Generation relative to the other three generations for age of initiation, cigarettes per day, smoking cessation, high cholesterol, and educational attainment. The 1964 Surgeon General’s report, which was approximately when baby boomers were entering the critical age window for SI, marked a pivotal shift in public awareness of smoking-related health risks. This abrupt policy shift triggered the beginning of an extended cultural re-evaluation of smoking norms. In earlier generations, when smoking was common, the social pressure to not smoke was minimal and individual characteristics were modestly associated with SI. As smoking became more stigmatized, we observe increasing magnitudes of the genetic correlations for people with heritable propensities for risk tolerance and impulsivity. A similar pattern emerges for females, where gendered norms have tended to discourage smoking in women. Thus, the distinct generational and gendered norms may have enabled or constrained SI as a function of other heritable traits, though likely via different heterogeneous associations implicating alternative biological mechanisms (**Figure 2**). These findings suggest that as smoking prevention policies and changing social norms discourage initiation, those who continue to smoke may increasingly be individuals with stronger underlying propensities toward risk taking and other behavioral characteristics associated with SI. Consequently, the success of earlier prevention efforts may change the composition of the population susceptible to initiation, highlighting the importance of adapting subsequent interventions to emerging patterns of risk. Changing patterns of initiation can also be seen in the genetic correlations between SI and social determinants of health. For example, in both females and younger generations, SI is associated with more neighborhood deprivation and lower rates of religious attendance, and for younger generations, less satisfaction with their lives. These findings suggest that the social and psychological characteristics associated with smoking initiation have become increasingly intertwined as smoking has become less socially acceptable. From a prevention perspective, this underscores the importance of addressing the broader social conditions associated with smoking risk, rather than focusing exclusively on tobacco use itself.

Overwhelmingly, genomic analyses have been conducted in European-like ancestry samples, limiting the generalizability and applicability to non-European populations.^20,21^ The current results emphasize the substantial overlap in the genetic architecture of SI across European, East Asian, Latino, and African American ancestry groups, both for sexes and the distinct generational birth cohorts. The substantial cross-ancestry concordance represents an important contribution to the understanding of SI. While we observe heterogeneity across sex and generations, we observe limited heterogeneity across ancestries. Accordingly, these findings support the assertion that multi-ancestry meta-analyses increased statistical power and expanded discovery beyond predominantly European-ancestry samples.^8^ However, similarity in genetic associations across ancestry groups should not be interpreted as evidence that ancestry or ancestry-associated social environments are irrelevant to smoking. Genetic ancestry is not a direct measure of social context, and differences in sample composition and statistical precision limit conclusions about ancestry-specific moderation.

The current analyses should be interpreted in light of several limitations. First, the fact that females and younger generations have higher levels of h^2^_snp_ does not imply the same underlying variants are being moderated in both cases. Sex and birth cohort likely moderate different genetic pathways to smoking initiation, as can be seen in the Manhattan plot (**Figure 2**). Second, detecting moderation for individual variants is underpowered, even with the unprecedented magnitude of the current sample size. As such, most moderated associations remain unidentified. The h^2^_snp_ results, by contrast, have substantially more power, and suggest moderation is relatively important. However, heritability analyses cannot provide a locus-level of biological specificity. Third, the generation moderation was conducted by binning individuals into age bins rather than treating age as a continuous variable. Any type of stratification decreases the power and specificity of the associations. Finally, while we cannot rule out survival bias as an explanation of SNP-by-generation interactions entirely, the broad pattern of results is only partially consistent with such explanations. Specifically, if people with a genetic predisposition to start smoking have a greater risk of mortality, we would expect a corresponding reduction in the magnitude of the genetic associations in the older generations, along with changes in allele frequencies that are too stark to be genetic drift. As such, survival bias would not explain situations where genetic associations are stronger in older generations. Further, the magnitude of survival bias for individual variants is relatively small, and likely smaller than the pattern of associations that we see in the current analyses. We use a simulation exercise with a strong assumption to test this hypothesis in **Supplementary Note G** and **Figure S13**.

GxE interactions have been difficult to identify in molecular data.^22^ Integrating GWAS meta-analyses with genome-wide heritability, genetic correlations, locus-level heterogeneity, and fine-mapping analyses allow us to identify consistent patterns of moderation for SI. Nevertheless, subgroup differences may also reflect differences in statistical power, linkage disequilibrium, and fine-mapping uncertainty, rather than genuinely distinct causal variants. Accordingly, these results identify promising targets for further investigation rather than establishing subgroup-specific biological mechanisms. Smoking provides a compelling case for examining gene–environment interplay because its social context has transformed dramatically over the past century, shifting from widespread acceptance to increasing stigmatization. Our findings demonstrate that sex and generation shape the genetic architecture of SI, highlighting how social context may influence the expression of genetic predispositions. More broadly, our findings challenge the notion that the genetic architecture of complex behaviors is universal or fixed, demonstrating that social transformations can reshape how genetic factors relate to complex behavioral outcomes. Yet sex and generation represent only two of many contextual forces that may shape genetic associations. For smoking prevention, these findings suggest that successful policies may reshape the population remaining susceptible to initiation. Accordingly, this requires prevention strategies to evolve alongside changing patterns of behavioral and social risk, targeting the emergent subgroups that resist smoking prevention efforts. Future research integrating measured policy environments, longitudinal behavioral data, and genetic information could help identify how interventions alter these patterns and inform more responsive approaches to tobacco prevention.

## Methods

### Overview of the Sample

We use the GWAS summary statistics for SI generated by the 23andMe team on August 9, 2024. To minimize the potential impact of population stratification, the summary statistics are provided separately for four genetic ancestry groups: European , African American, East Asian, and Latino (see **Supplementary Table S1** for the precise sample sizes for each ancestry). For each ancestry group, the 23andMe team provided one full set of GWAS summary statistics as well as 23 additional GWAS summary statistics stratified by sex (2 groups for each ancestry), geographic US region (5 groups for each ancestry), and birth year cohort (16 groups run in 5-year birth year bins for each ancestry), allowing us to investigate well-powered gene-by-environment (GxE) interactions and multi-ancestry analyses. Sample sizes for each of these groups is included in **Supplementary Table S1**.

### Phenotype Definition

Smoking Initiation (SI) is a binary phenotype with a value of 1 if an individual self-reported having ever regularly smoked or having used at least 100 cigarettes in their lifetime and 0 otherwise. The numbers of cases and controls for each of our samples and subsamples are reported in **Supplementary Table S1**.

### Sample Inclusion Criteria

GWAS participants were 23andMe research participants aged 18 or older who provided informed consent for use of their fully de-identified data for research. They also had genome-wide genotype data and complete information on the SI phenotype, age, sex, and principal components of the genetic variance-covariance matrix (the first five for European and East Asian ancestry, the first six for African American ancestry, and the first nine for Latino ancestry). For each analysis, the sample was restricted to unrelated individuals from one of four ancestry groups (European, African American, East Asian, or Latino). In the stratified analyses, samples were screened to ensure only one family member is included per subsample. However, to ensure maximum statistical power, multiple family members were allowed to contribute to different sex or birth cohort analyses. Accordingly, sample size varies across the stratified analyses. Furthermore, for the analyses stratified by region, the sample was further restricted to individuals residing in the US.

Detailed inclusion criteria, including genetic inclusion criteria, can be found in **Box 1**. The sample size differs slightly across analyses, reflecting variation in the availability of research participants who met all relevant inclusion criteria and data requirements for a given analysis (**Box 1**).

#### Box 1.

**GWAS participant inclusion criteria (23andMe Research cohort)**

| Participants included in the GWAS meet the following criteria: |
|---|
| 1. Provided informed consent to participate in the 23andMe Research.<br>2. Successfully genotyped across the genome. |
| Demographics and required covariates:<br>3. Aged 18 years or older.<br>4. Have available data on the phenotype, age (as defined by 5-year birth year bins spanning 1924-2003), sex (female and male), and ancestry-specific principal components (the first five for European and East Asian ancestry, the first six for African American ancestry, and the first nine for Latino ancestry). |
| Biological Sex:<br>a. Female<br>b. Male |
| Generation / Birth Cohort (as defined by 5-year birth year bins spanning 1924-2003):<br>a) Silent Generation (1924 - 1943)<br>b) Baby Boomers (1944 - 1963)<br>c) Generation X (1964 - 1983)<br>d) Millennials (1984 - 2003)<br>*The names for these generations were given according to the largest percentage of individuals in each bin. Because we meta-analyzed GWAS summary statistics from four 5-year birth cohort bins to derive each of these four larger generations, the bins did not fit cleanly into the standard birth years for these generations (usually observed as Silent Generation: 1928–1945; Baby Boomers: 1946–1964; Generation X: 1965–1980; Millennials: 1981-1996; Generation Z: 1997–2012). Therefore, we named the generations according to the largest portion represented in each bin, recognizing that there is an imperfect match. |
| Geography (applied only in the samples stratified by region):<br>5. Reside in one of the five 23andMe defined US regions:<br>a. The *Northeast* region includes Maine, Massachusetts, Rhode Island, Connecticut, New Hampshire, Vermont, New York, Pennsylvania, New Jersey, Delaware, and Maryland.<br>b. The *Southeast* region includes West Virginia, Virginia, Kentucky, Tennessee, North Carolina, South Carolina, Georgia, Alabama, Mississippi, Arkansas, Louisiana, and Florida.<br>c. The *Midwest* region includes Ohio, Indiana, Michigan, Illinois, Missouri, Wisconsin, Minnesota, Iowa, Kansas, Nebraska, South Dakota, and North Dakota.<br>d. The *Southwest* region includes Texas, Oklahoma, New Mexico, and Arizona.<br>e. The *West* region includes Colorado, Wyoming, Montana, Idaho, Washington, Oregon, Utah, Nevada, California, Alaska, and Hawaii. |
| Genetic ancestry group and Relatedness filtering:<br>6. Belong to one of the four 23andMe defined ancestry groups: European, African American, East Asian, or Latino.<br>7. Are unrelated within the dataset. Participants are considered related if they share more than 700 centimorgans (cM) of identity-by-descent (IBD), including regions where they share one or both genomic segments IBD. |

### 23andMe Ancestry Classifier

23andMe ancestry classifier algorithm determines participant ancestries through an analysis of local ancestry. It first partitions phased genotyped data into short windows of about 300 SNPs. Within each window, we use a support vector machine (SVM) to classify individual haplotypes into one of 45 worldwide reference populations. The SVM classifications are then fed into a hidden Markov model (HMM) that accounts for switch errors and incorrect assignments, and gives probabilities for each reference population in each window. Finally, we used simulated admixed individuals to recalibrate the HMM probabilities so that the reported assignments are consistent with the simulated admixture proportions. For variant imputation, we are aggregating the probabilities of the 45 reference populations into five main ancestries:

#### Box 2.

**23andMe ancestry classifier**

| **Ancestry** | **Classification Criteria** |
|---|---|
| European | European + Middle Eastern > 0.97, European > 0.90 |
| East Asian | Japanese Korean + Chinese Southeast Asian + Manchurian Mongolian > 0.94 |
| South Asian | South Asian > 0.97 |
| Middle Eastern (& North African) | Middle Eastern + European > 0.97, Middle Eastern > 0.90 |
| African American + Latinos | European + African +East Asian + Native American + Middle Eastern > 0.90, African + Native American > 0.01 |

African Americans and Latinos are admixed with varying contributions from Europe, Africa and the Americas. Therefore, no single threshold of genome-wide ancestry will be able to effectively separate African Americans and Latinos. However, the distributions of the length of segments of European, African and American ancestry are very different between African Americans and Latinos, because of distinct admixture timing between the three ancestral populations in the two ethnic groups. Therefore, we trained a logistic classifier that takes one participant’s length histogram of segments of African, European and American ancestry, and predicts whether the participant is likely African American or Latino.

A detailed description of 23andMe’s local ancestry classifier can be found here: https://www.23andme.com/ancestry-composition-guide/

### Genotyping and Imputation

Individuals in the sample were genotyped using one of the five 23andMe team genotyping platforms. Versions 1 and 2 platforms used the Illumina HumanHap550 and BeadChip. The Version 3 platform was based on the Illumina OmniExpress and BeadChip. Versions 4 and 5 platforms utilized the Illumina custom array and the Illumina Infinium Global Screening Array, respectively. The genotype imputation was conducted using 23andMe’s r7 imputation panel, which integrates data from three sources: the publicly available Human Reference Consortium (HRC), the UK BioBank (UKBB) 200K Whole Exome Sequencing (WES) reference panels, and the 23andMe internal reference panel. All studies used Beagle 5 for imputation^23^. Roughly 99.7 million variants were imputed for each research participant after determining genetic ancestry and phasing high quality genotyped variants within each ancestry. Because there were five genotyping platforms used, imputation was performed independently for each platform.

### Quality Control of Genetic Data

The 23andMe team applies quality control (QC) procedures independently to genotyped and imputed variants. For the genotyped variants, genetic association tests are conducted using all available genotypes across all the five genotyping platforms. After the association testing, SNPs are excluded if they: (1) are genotyped only on platform v1 and/or v2 due to small sample size; (2) are located on chromosome M and Y, which are currently considered unreliable; (3) fail a parent-offspring transmission test; (4) have a genotype call rate below 90%; (5) show evidence of genotype date effects; (6) exhibit a large sex effect; (7) map to multiple genomic positions in the reference genome; (8) have MAF equals 0% (MAF=0); or (9) have Hardy Weinberg Equilibrium (HWE) with a *p*-value greater than 1e-20 (HWE>1e-20). For imputed variants, we first evaluate the global quality of a specific variant imputation in each genotyping platform: if the imputation R^2^<0.3 for a specific variant in one of the genotyping platforms, the imputation dosages are masked and are not included in the association test. Because the number of participants genotyped in each five genotyping platforms is very different (>90% of participants were genotyped on v4 or v5), the effective number of samples for each variant can be highly variable. To limit the difference in effective number of samples between variants, we completely mask variants that are not imputed correctly in either v4 or v5. Following association testing, imputed SNPs are excluded if they: (1) have imputation quality (R^2^) less than 0.5 (R^2^<0.5), (2) show evidence of imputation batch effects; (3) exhibit large sex effects; or (4) have MAF equals 0% (MAF=0). Finally, across both genotyped and imputed variants, SNPs are excluded if the number of individuals contributing data for the SNP is less than 20% of the total GWAS sample size, or if the logistic regression fails to converge due to complete separation, as indicated by absolute log-odds ratio or standard errors greater than 10. Additionally, we further excluded SNPs with MAF less than 1% (MAF < 0.01) in our analysis, because single variant tests are underpowered for rare variants.

### Genetic Association Testing

The genetic association testing in the full samples for each of the 4 ancestries was performed using a logistic regression model assuming additive allelic effects among unrelated individuals:

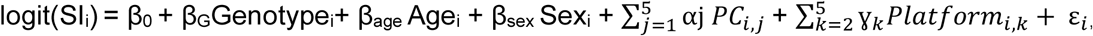

where SI_i_ is the binary Smoking Initiation phenotype for individual *i*. Genotype_i_ represents the individual’s genotype at a specific basepair in the genome, Age_i_ and Sex_i_ are covariates for age and sex, PC_i,j_ indicates the inclusion of ancestry-specific principal components (the first five for European and East Asian ancestry, the first six for African American ancestry, and the first nine for Latino ancestry), and Platform_i,k_ is an indicator variable for genotyping platforms used (*k*), accounting for genotype batch effects.

GWAS were only conducted for each subsample split (ancestry, sex, US region, and birth year bin) when there were at least 100 SI cases for each of the 24 possible subsamples for each ancestry group. For this reason, even though there were large enough SI cases in the full Middle Eastern and South Asian ancestry groups, these results were not included, because the stratified subsamples within these ancestry groups were too small which could produce excessive false positive or false negative associations.

When GWAS were run for stratified groups within ancestries, covariates were excluded where appropriate. For example, sex was not included as a covariate in the GWAS regression when analyses were stratified by sex. Birth year was still included as a covariate in birth year-bin stratified GWAS, since there was still age-specific variation with birth year bins.

### Genomic Meta-Analyses

We conducted sequential meta-analyses across subgroups within each moderator using inverse variance weighting implemented in METAL^16^ for sex and generation. To simplify the generation analyses and to minimize the impact of population stratification, within each ancestry subsample, we meta-analyzed the 5-year birth year bins into four generations. Then, we meta-analyzed the specific levels of the moderators (sex: females and males; generations: Silent Generation, Baby Boomers, Generation X, and Millennials; geographic region: North East, South East, Midwest, South West, and West) across ancestry groups. Finally, we meta-analyzed the summary statistics across different levels of the same moderator. At each stage, we evaluated whether SNP effect sizes differ systematically across subgroups using the p-value from Cochran’s Q test for heterogeneity.

### Cross-Ancestry Genetic Correlation

We estimate cross-ancestry genetic effect correlations using the Popcorn software package.^17^ Popcorn uses GWAS summary statistics and ancestry-specific linkage disequilibrium reference panels to estimate the correlation of SNP effect sizes across ancestry groups at variants common to both populations. We used ancestry-specific reference panels from the 1000 Genomes Project.^24^ Pairwise analyses are conducted for all ancestry combinations within each moderator group. To assess the extent of shared genetic architecture across populations, we estimate the genetic effect correlation (*ρ_e_*) and test whether the correlation differs significantly from one.

### Testing for Heterogeneous SNPs for Moderation

We used a two-step criteria to assess gene-by-environment (GxE) interactions. First, the variant had to have a significant heterogeneity p-value (*HetPval*) in the meta-analysis across different levels of a moderator (e.g., females vs males) and at least one level of the moderator had to be significant (e.g., males or females). We used the standard genome-wide significance threshold of *P* < 5 × 10^−8^for both tests to account for multiple testing. While this two-step criteria is extremely conservative, it enhances the robustness of the moderated genetic associations.

### Functional Mapping and Annotation (FUMA) of GWAS

To identify independent significant SNPs, lead SNPs, genomic risk loci, and mapped genes, we use the SNP2GENE module in FUMA^25^. Independent significant SNPs are defined as variants surpassing the genome-wide significance threshold (*P* < 5 × 10^−8^) and are mutually independent at an LD threshold of *r*^2^ ≥ 0.6. Lead SNPs were then identified by applying an LD filter of *r*^2^ ≥ 0.1, with each lead SNP serving as the sentinel variant for an association signal. FUMA aggregates nearby LD blocks surrounding lead SNPs into broader genomic risk loci by merging regions located within 250kb of one another. Each locus may contain one or more lead SNPs, together with additional SNPs in linkage disequilibrium, thereby capturing the local genetic architecture of each association signal. To identify genomic risk loci and mapped genes associated with heterogeneous variants, we use *P*-values derived from the heterogeneity statistics obtained from the meta-analyses.

### Autoregressive Latent Change Score Model

To investigate whether the genetic overlap between SI and external phenotypes varies across generations, we estimated an autoregressive latent change score model (LCSM) within the Genomic SEM framework^15^ using GWAS of SI for the Silent Generation, Baby Boomers, Generation X, and Millennials as input. Latent innovation factors represent the newly emerging genetic variance in SI within each successive generation after accounting for the genetic liability carried over from the immediately preceding generation.

Autoregressive paths capture the transmission of genetic influences from one generation to the next. We examined 35 external phenotypes spanning three domains: substance use, sociodemographic characteristics, and psychiatric and medical conditions. Statistical significance is evaluated using both nominal (*p* < 0.05) and Bonferroni-corrected thresholds accounting for all phenotype-by-innovation tests. For each external phenotype, we estimate its genetic correlation with each generation-specific innovation factor. By correlating each phenotype with the generation-specific innovation factors, this approach examines whether the genetic overlap between SI and each phenotype is generation-specific, thereby identifying the generations during which associations with external traits are most strongly associated with newly emerging genetic liability to SI. Models are estimated using Genomic SEM with diagonally weighted least squares (DWLS) based on the linkage disequilibrium score regression (LDSC)-derived genetic covariance matrix and sampling covariance matrix.

As a complementary analysis, we estimate a two-generation LCSM including only the earliest (Silent Generation) and latest (Millennials) generations. The resulting innovation factor captures genetic influences on SI in the Millennials that were not explained by genetic liability in the Silent Generation. We estimate genetic correlations between this innovation factor and the same set of external phenotypes to evaluate whether comparing genetically more distant generations yields even stronger evidence of generation-specific genetic overlap.

### Generalized Least Squares Regression

As a complementary approach to assessing temporal heterogeneity in genetic effects, we examine linear trends in SNP effect sizes across 16 birth-year cohorts. For each lead SNP identified using heterogeneity statistics within the 1,580 main effect and 8 moderated effect genomic risk loci, we estimate a generalized least squares (GLS) model in which the SNP effect size is regressed on an ordered birth-year-cohort index, with observations weighted by the inverse of the variance of the estimated SNP effect. Effect alleles are harmonized such that positive effect sizes consistently represent the risk-increasing direction. The estimated GLS slope therefore captures the average linear change in the risk-increasing effect across consecutive birth-year cohorts, with positive slopes indicating strengthening genetic effects in more recent birth cohorts and negative slopes indicating weakening effects. Statistical significance is determined using Bonferroni-corrected thresholds based on the number of SNPs tested within each set.

### Fine-Mapping Analysis

We performed statistical fine-mapping of genome-wide significant loci identified in the cross-ancestry meta-analyses using the Sum of Single Effects (SuSiE) method. ^18,19^ SuSiE uses GWAS summary statistics^19^ and linkage disequilibrium (LD) information to estimate posterior inclusion probabilities (PIPs) for variants within each locus, thereby quantifying the likelihood that a variant is causally associated with the trait. Because we only have GWAS summary statistics data, we used ancestry-specific reference panels from the 1000 Genomes Project^24^ to calculate LD information. Variants with PIP > 0.8 were classified as high-confidence (high-PIP) candidate causal variants, and loci containing at least one such variant were classified as high-PIP loci. Fine-mapping results were subsequently compared across sex and generational strata to evaluate the consistency of prioritized variants and loci underlying SI.

### European Ancestry Sample Description

Because the European ancestry group exhibits well-characterized linkage disequilibrium (LS) structure, is less admixed than the other ancestry groups, and constitutes the best powered sample among all genetic ancestries, comprising 2,217,534 cases and 3,252,222 controls (**Table S1**), we focus our Linkage Disequilibrium Score Regression analyses on this population. We use GWAS summary statistics for this sample stratified by sex (N_female_ = 2,440,320; N_male_ = 2,016,119), by 5-year birth year bins, also aggregated into 4 generations through meta-analysis (N_1924-1943_ = 366,023; N_1944-1963_ = 1,677,905; N_1964-1983_ = 1,826,054; N_1984-2003_ = 1,599,774) (**Table S1**). These summary statistics represent the largest single-sample US-based genetic database for SI, both overall and when split by sex, five US geographic regions, or birth cohort.

### Heritability and Genetic Correlation

We used Linkage Disequilibrium Score Regression (LDSC) in the European-like ancestry subsample to estimate (1) the SNP heritability of SI, (2) the genetic correlations across different levels of the sex-stratified meta-analyses (males vs females) and generation-stratified analyses (1924-1943, 1944-1963, 1964-1983, vs 1984-2003 birth cohorts), and 3) to estimate cross-trait genetic correlations between SI and a set of curated phenotypes (e.g., educational attainment or body mass index) using publicly available GWAS summary statistics.^26,4,27–34^ As LD patterns vary across ancestry groups, LDSC cannot be applied to pan-ancestry meta-analytic summary statistics. Therefore, the LDSC analyses were restricted to the European subsample. The European-like subsample was the largest homogeneous group, and most available, relevant phenotypes used in the cross-trait analyses focus on EUR-like ancestry groups. As such we used linkage disequilibrium reference panels from the 1000 Genomes Project^24^ for our LDSC reference panel^35^, using only HapMap3 SNPs.^36^ We tested pairwise equality tests of genetic correlation across sex and birth cohort subgroups for all phenotypes.

### Ethics Statement

The 23andMe data was collected in compliance with all applicable laws, from human subjects who provided informed consent under an IRB-approved protocol in accordance with US federal guidelines for Protection of Human Subjects. Participants provided informed consent and volunteered to participate in the research online, under a protocol approved by the external AAHRPP-accredited Salus IRB (https://www.versiticlinicaltrials.org/about-us/our-brands/salus-irb). 23andMe has complied and will comply with all applicable laws and ethical requirements to protect the identity and privacy of human subjects from whom the data was collected. At Purdue University, the following determination was made after IRB review under IRB study number STUDY2026-00001158: “The Purdue Human Research Protection Program (HRPP) Office determined that the proposed activity is not research involving human subjects as defined by DHHS and FDA regulations.” At Purdue University, the executed legal Data Transfer Agreement (DTA) with 23andMe is maintained through Legal Counsel under agreement DATA25036938.

## Data Availability

The summary statistics for our main effect and moderated effect loci and variants are available in **Table S13**, which is included as part of our **Supplemental Tables**. We are limited to providing only up to 10,000 SNPs due to contractual agreements with 23andMe Research Institute based on the privacy policies restricting the release of summary statistics. Access to the full summary statistics of the 23andMe sample (for all SNPs) can be obtained by qualified researchers through a data transfer agreement with 23andMe Research Institute that protects the privacy of the 23andMe participants. Researchers interested in the full meta-analysis summary statistics containing 23andMe data must apply to 23andMe. Please visit https://research.23andme.org/dataset-access for more information and to apply to access the data.

## Code Availability

The code for this paper is available through GitHub (https://github.com/yjeong0413/GxE-of-Smoking-Initiation.git).

## Acknowledgements

We would like to thank the research participants and employees of 23andMe Research Institute for making this work possible.

